# Ejection fraction on a budget: mapping the accuracy-compute trade space for video-based ejection fraction estimation

**DOI:** 10.64898/2026.09.02.26362055

**Authors:** Aryan Pandey, Kushaan Sharma, Aryan Shah

## Abstract

Deep video networks estimate left ventricular ejection fraction (EF) from echocardiograms with expert-level accuracy, but the compute cost of running them is rarely reported, which leaves anyone building a handheld or bedside tool without guidance on what to deploy. We measured the accuracy versus compute trade space for EF estimation on EchoNet-Dynamic by training 22 configurations that vary clip length (8 to 64 frames), frame sampling period (1 to 4), and backbone (R(2+1)D-18, R3D-18, MC3-18, X3D-S, X3D-M, and a 2D ResNet-18 with temporal pooling), under one fixed training recipe. Every configuration was scored on accuracy (mean absolute error, *R*^2^, Bland-Altman agreement), on clinical utility (sensitivity and specificity at the EF 40% and 50% treatment thresholds, error stratified by EF band), and on cost (floating point operations, parameters, GPU and CPU latency, peak memory) under a single frozen measurement protocol. Headline claims were stress tested with replicate training seeds. Sparse temporal sampling consistently beat dense sampling: at a fixed frame count, period 4 improved mean absolute error by about one full point over period 1 across all nine cross-seed pairings while also cutting per-video cost. A plain R3D-18 achieved the best point accuracy in the study (mean absolute error 3.99), statistically tied with the reference, at 19% less CPU latency, and a 16-frame, period-4 R(2+1)D-18 halved the reference cost with no statistically confirmed accuracy loss, though with a small seed-consistent disadvantage that no single seed reveals. Removing temporal modeling entirely collapsed accuracy (mean absolute error 5.65), placing a floor under how cheap this task can get. We release the code, the cost protocol, and all per-configuration results.

## Introduction

Left ventricular ejection fraction is the fraction of blood the left ventricle expels per beat. It is the single most consequential number in a routine echocardiogram: heart failure is classified by it, and treatment decisions hinge on whether a patient sits below 40% or below 50% [1–3]. Guideline practice estimates EF from manual endocardial tracings at end-diastole and end-systole, most commonly by the biplane method of disks [4], a procedure that is operator-dependent: reported interobserver variation spans 7.6 to 13.9 percent [5], and even across imaging modalities EF estimates for the same patient disagree substantially [6].

Deep learning has attacked this measurement problem along two lines. The first automates the guideline procedure itself: segmentation networks delineate the left ventricle frame by frame, and EF follows from the segmented end-diastolic and end-systolic volumes. This line runs through the CAMUS benchmark [7] and remains active, including recent attention-augmented U-Net variants [8] and heuristic-corrected segmentation pipelines [9] evaluated on the same dataset used here. The second line regresses EF directly from the video. Ouyang et al. [5] showed that a spatiotemporal convolutional network trained end-to-end on the EchoNet-Dynamic dataset estimates EF from apical four-chamber video with a mean absolute error (MAE) of 4.1 points, inside the human interobserver range, and a blinded randomized trial subsequently found that cardiologists could not distinguish this model’s initial EF assessments from sonographer measurements and corrected them less often [10]. Direct regression has since been extended with video transformers [11, 12], graph neural networks over sampled frames [13], and pediatric cohorts [14]. Video-based EF estimation is, by any measure, one of the most credible near-term applications of deep learning in cardiology.

Accuracy, however, is only half of a deployment decision. The published EchoNet-Dynamic model processes 32-frame clips through an R(2+1)D-18 backbone, and the natural targets for automated EF, handheld probes, point-of-care ultrasound carts, and screening workflows in settings without a reading cardiologist, are exactly the environments where compute, memory, and battery are scarce. Handheld ultrasound devices already put diagnostic imaging in coat pockets, and their computational budgets are a fraction of a workstation’s [15, 16]. The original authors swept clip lengths, sampling periods, and backbones and reported how accuracy responded [5], but no cost was attached to any of those points, and to our knowledge nobody has since assembled the accuracy and the cost of this task systematically under one protocol. Later work has produced individual efficient or compact architectures for EF estimation [12, 17, 18], and resource-efficient echocardiography models have been demonstrated on edge hardware for related tasks [19], which answers whether an efficient model can be built, but not what the whole trade space looks like: how much accuracy each unit of compute buys, where the returns flatten, and where accuracy collapses. Model-compression techniques such as pruning, quantization, and distillation [20–22] attack the same problem from an orthogonal direction; they can be applied on top of any architecture, but they do not answer which architecture and input configuration to start from.

This paper maps that trade space. We trained 22 configurations spanning the three axes that drive inference cost, clip length, temporal sampling period, and backbone architecture, under one fixed training recipe on EchoNet-Dynamic, and paired every accuracy measurement with a full cost profile taken under a single frozen protocol. Three design choices distinguish this from the sweep in the original paper. First, cost is measured, not implied: floating point operations (FLOPs), parameters, GPU and CPU latency, peak memory, and model size are recorded for every configuration under identical conditions, including the per-video cost that follows from how many clips each configuration needs to tile a real video. Second, the backbone axis includes architectures that postdate the original work, X3D [23] and a 2D-network-with-pooling floor, which are what a deployment team would consider today. Third, clinical error structure is reported at every operating point, not only for the winning model: sensitivity and specificity at the 40% and 50% thresholds and MAE stratified by EF band, because a two-point error at EF 25 and a two-point error at EF 41 have very different consequences.

We also stress tested the claims that emerged. The comparisons a reviewer would most likely question, the cheap configuration that appears to match the reference, and the finding that sparse sampling beats dense sampling, were re-run under two additional training seeds that were pre-registered before any replicate training began.

The result is a Pareto frontier with three practical recommendations at different budgets, one robust and previously unquantified finding about temporal sampling, and a measured compute floor below which accuracy falls apart. All code, configurations, per-video predictions, and the cost protocol are released.

## Materials and methods

### Dataset and ethics

EchoNet-Dynamic [5] contains 10,030 apical four-chamber echocardiogram videos from distinct patients at Stanford Medicine (2016–2018), each de-identified, cropped, masked, and downsampled to 112 *×* 112 pixels, with an expert-measured EF label. We used the patient-level train, validation, and test split that ships with the dataset: 7,465 training, 1,288 validation, and 1,277 test videos. Test-set EF bands contain 83 videos below 30%, 77 between 30 and 40, 241 between 40 and 55, and 876 at or above 55; 160 test videos have EF below 40% and 285 below 50%. Data were obtained under the Stanford research use agreement, and no videos, frames, or derived imagery are redistributed. Input normalization used per-channel mean and standard deviation computed once on the training split. This work is a secondary analysis of a fully de-identified, publicly available research dataset, so no additional institutional review board approval was required.

### Configurations

Each configuration is defined by a backbone, a clip length *F* (frames per forward pass), and a sampling period *P* (stride between sampled frames), so one clip spans *F × P* raw frames of video. Stage one fixed the backbone to the reference R(2+1)D-18 [24] and crossed *F ∈ {*8, 16, 32, 64} with *P ∈ {*1, 2, 4} (12 configurations, including the reference setting of 32 frames at period 2). Stage two fixed the two most informative clip settings from stage one, the reference geometry (32 *×* 2) and the cheapest setting that matched the best stage-one accuracy within half a point (16 *×* 4), and varied the backbone across the five architectures described next, for 10 further configurations.

### Backbone architectures

All backbones are convolutional; each ends in global spatiotemporal average pooling followed by a single linear unit that regresses EF in percent directly, with no output nonlinearity. Direct 3D convolution over video was established by C3D [25] and matured through two-stream inflated networks [26]; the residual variants used here follow Tran et al. [24].

*R(2+1)D-18* (31.3 million parameters; 81.3 GFLOPs per 16-frame clip) is the reference backbone from the EchoNet-Dynamic study [5]. It follows the 18-layer residual topology [27] but factorizes every *k*_*t*_ *× k × k* spatiotemporal convolution into a 1 *× k × k* spatial convolution followed by a *k*_*t*_ *×* 1 *×* 1 temporal convolution, with the intermediate channel width chosen so the factorized pair matches the parameter count of the full 3D kernel; the factorization doubles the number of nonlinearities per block [24].

*R3D-18* (33.2 million parameters; 81.5 GFLOPs per 16-frame clip) is the unfactorized counterpart: the same 18-layer residual topology with full 3 *×* 3 *×* 3 convolutions throughout [24].

*MC3-18* (11.5 million parameters; 86.8 GFLOPs per 16-frame clip) uses mixed convolutions: 3D convolutions in the early residual groups, where motion information is extracted, and 2D convolutions in the later groups [24].

*X3D-S and X3D-M* (3.0 million parameters each; 5.0 and 9.8 GFLOPs per 16-frame clip including the internal upsampling described below) are mobile-scale video networks obtained by progressively expanding a tiny 2D image architecture along frame-rate, resolution, width, and depth axes, with channelwise-separable convolutions carrying most of the computation [23]. They are the most parameter- and FLOP-efficient video backbones with maintained Kinetics-400 pretrained weights available at the time of this study; MoViNets [28] occupy a similar design point but lacked a maintained PyTorch port with pretrained weights and were excluded.

*2D ResNet-18 with temporal pooling* (11.2 million parameters; 15.6 GFLOPs per 16-frame clip) applies an ImageNet-pretrained 2D ResNet-18 [27, 29] to each frame independently, mean-pools the resulting 512-dimensional feature vectors over time, and regresses EF from the pooled feature. Because pooling discards frame order, this model sees the same pixels as the video backbones but cannot represent motion; it deliberately serves as the temporal-modeling floor of the study.

All 3D backbones were initialized from Kinetics-400 pretraining [30]; the 2D backbone from ImageNet. X3D requires one documented deviation. Its downsampling schedule is built for 160-pixel (X3D-S) or 224-pixel (X3D-M) input, and at this dataset’s native 112 pixels a deep feature map shrinks below its own convolution kernel, which is a hard runtime failure rather than a soft accuracy question. We therefore upsample the decoded 112-pixel clip bilinearly to the native size inside the model, immediately before the first X3D layer. The decode pipeline is untouched and identical for every configuration, and the upsampling cost is included in X3D’s measured latency and FLOPs. The consequence is a confound we cannot remove: X3D’s pretrained weights expect sharp native-resolution video, and an upsampled 112-pixel source has the right shape but not the same detail. X3D’s accuracy here therefore cannot be cleanly attributed to the architecture as opposed to the degraded input, and every X3D number in this paper carries that caveat.

### Training

Every run used the same recipe, so configurations differ only along the swept axes. Let *f*_*θ*_ denote a backbone with parameters *θ* and *x*_*i*_ *∈* ℝ ^3*×F ×*112*×*112^ a sampled clip with label *y*_*i*_ (EF in percent). Training minimizes the mean squared error over each mini-batch of size *B*,

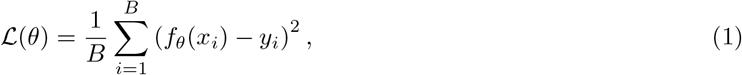

by stochastic gradient descent with learning rate 10^*−*4^, momentum 0.9, and weight decay 10^*−*4^, for 30 epochs with the learning rate multiplied by 0.1 every 10 epochs. The effective batch size is 20 for every run; where a configuration’s activation memory exceeds the GPU, the physical batch is reduced to a divisor of 20 and gradients are accumulated across micro-batches before each optimizer step, so the update itself is identical across the study. Training uses fp16 mixed precision with dynamic loss scaling. Each epoch draws one clip per training video at a uniformly random start index; spatial augmentation follows the EchoNet recipe of zero-padding 12 pixels on each side and taking a random 112 *×* 112 crop, equivalent to a random translation of up to *±*12 pixels. After each epoch the model is evaluated on the validation split (deterministic first clip per video), and the checkpoint with the lowest validation MAE is carried forward to testing. The grid uses a fixed seed (20260814) for all random number generators. The recipe is a shortened version of the original EchoNet schedule (45 epochs, learning rate step at 15) [5] adapted to a consumer GPU; the baseline subsection of the Results verifies that it reproduces the published accuracy. All experiments ran in Python 3.11 with PyTorch 2.6.0 (CUDA 12.4) and torchvision [31] on a single NVIDIA RTX 2070 Super (8 GB); the full 30-run study, including seed replicates, took 83 GPU hours.

### Test protocol

For test-set inference, each video of *T* raw frames is tiled into

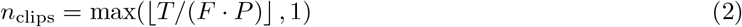

non-overlapping clips starting at frame zero (videos shorter than one span are loop-padded), and the video-level prediction is the mean of the per-clip predictions. This matches the test-time behavior of the original EchoNet repository and is used for every accuracy number reported here.

### Cost protocol

Cost was measured once per configuration under a frozen protocol on a single machine, and no number measured under different conditions appears in any table. FLOPs were counted with fvcore [32] (reported as 2*×* multiply-accumulates), with unsupported operators logged rather than silently dropped. Latency is the median of 100 timed forward passes after 20 discarded warmup passes, at batch size 1 in fp32 evaluation mode: on GPU with device synchronization around each pass, and on CPU restricted to 4 threads as a proxy for portable hardware. Peak GPU memory covers one inference forward pass, and model size is the fp32 checkpoint size.

Every input-dependent quantity is reported per clip and per video. The per-video figure multiplies per-clip cost by the mean number of non-overlapping clips needed to tile a test video at that geometry, which depends only on *F × P* and the real distribution of video lengths. This term matters: at 8 frames, a period-1 clip spans 8 raw frames and a test video needs 21.7 clips on average, while a period-4 clip spans 32 raw frames and needs 5.0, so per-video cost differs by a factor of four at identical per-clip cost. The primary cost axis throughout is CPU latency per video; FLOPs, GPU latency, and memory are reported alongside.

Measured cost excludes video decoding and preprocessing, which are deployment-specific; the limitation is discussed in the Discussion.

Two rows of Table 1 deserve a note. Per-clip latency depends only on the input tensor shape, so configurations that share a clip length should show the same per-clip latency, and most do, within about 3% on CPU. The R(2+1)D-18 32 *×* 4 and 64 *×* 4 rows measured 13% and 8% above the period-1 row of the same clip length, with wider interquartile ranges across the 100 timed runs (64 and 104 ms, against 22 and 27 ms for the other 32-frame rows and 43 and 88 ms for the other 64-frame rows), which points to background load on the shared measurement machine during those two runs rather than a protocol difference. Both rows are reported as measured; neither lies on the frontier, and no conclusion depends on them.

**Table 1.** Cost profile of every configuration, sorted by CPU latency per video. Measured under the frozen protocol (fp32, batch 1; CPU limited to 4 threads; median of 100 runs after 20 warmup runs). ^*†*^X3D rows: input upsampled from 112 pixels to the architecture’s native resolution inside the model; see Materials and methods.

| Configuration | GFLOPs<br>/clip | Params<br>(M) | Disk<br>(MB) | Clips<br>/video | CPU<br>(ms/video) | GPU<br>(ms/video) | Mem<br>(MB) |
| --- | --- | --- | --- | --- | --- | --- | --- |
| 2D ResNet-18 + pool 16f×p4 | 15.6 | 11.2 | 45 | 2.28 | 163 | 8.2 | 84 |
| X3D-S 16f×p4 <sup>†</sup> | 5.0 | 3.0 | 12 | 2.28 | 223 | 41.0 | 86 |
| 2D ResNet-18 + pool 32f×p2 | 31.1 | 11.2 | 45 | 2.28 | 327 | 11.8 | 113 |
| X3D-M 16f×p4 <sup>†</sup> | 9.8 | 3.0 | 12 | 2.28 | 490 | 41.0 | 145 |
| X3D-S 32f×p2 <sup>†</sup> | 10.0 | 3.0 | 12 | 2.28 | 506 | 42.7 | 148 |
| R3D-18 16f×p4 | 81.5 | 33.2 | 133 | 2.28 | 564 | 32.7 | 212 |
| R(2+1)D-18 16f×p4 | 81.3 | 31.3 | 125 | 2.28 | 688 | 43.0 | 235 |
| MC3-18 16f×p4 | 86.8 | 11.5 | 46 | 2.28 | 688 | 33.5 | 121 |
| R(2+1)D-18 32f×p4 | 162.6 | 31.3 | 125 | 1.07 | 731 | 38.2 | 335 |
| R(2+1)D-18 8f×p4 | 40.6 | 31.3 | 125 | 5.05 | 761 | 54.5 | 185 |
| X3D-M 32f×p2 <sup>†</sup> | 19.6 | 3.0 | 12 | 2.28 | 1103 | 48.9 | 266 |
| R3D-18 32f×p2 | 163.0 | 33.2 | 133 | 2.28 | 1122 | 59.1 | 278 |
| MC3-18 32f×p2 | 173.6 | 11.5 | 46 | 2.28 | 1339 | 62.1 | 188 |
| R(2+1)D-18 32f×p2 (ref.) | 162.6 | 31.3 | 125 | 2.28 | 1381 | 77.5 | 335 |
| R(2+1)D-18 64f×p4 | 325.2 | 31.3 | 125 | 1.00 | 1418 | 66.8 | 532 |
| R(2+1)D-18 64f×p2 | 325.2 | 31.3 | 125 | 1.07 | 1431 | 70.6 | 532 |
| R(2+1)D-18 16f×p2 | 81.3 | 31.3 | 125 | 5.05 | 1510 | 95.6 | 235 |
| R(2+1)D-18 8f×p2 | 40.6 | 31.3 | 125 | 10.61 | 1636 | 115.6 | 185 |
| R(2+1)D-18 64f×p1 | 325.2 | 31.3 | 125 | 2.28 | 2990 | 156.5 | 532 |
| R(2+1)D-18 32f×p1 | 162.6 | 31.3 | 125 | 5.05 | 3028 | 171.8 | 335 |
| R(2+1)D-18 16f×p1 | 81.3 | 31.3 | 125 | 10.61 | 3208 | 201.1 | 235 |
| R(2+1)D-18 8f×p1 | 40.6 | 31.3 | 125 | 21.73 | 3267 | 238.9 | 185 |

### Metrics and statistical analysis

For *n* test videos with reference values *y*_*v*_ and predictions *ŷ*_*v*_ (the per-video mean of clip predictions defined in the Test protocol), accuracy is summarized by

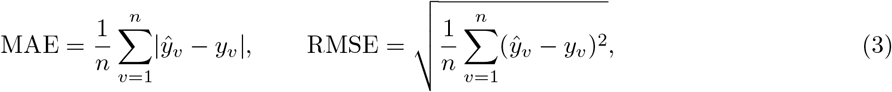

and the coefficient of determination

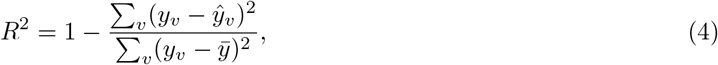

where 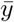 is the test-set mean. Agreement is additionally assessed by Bland-Altman analysis [33]: the bias is the mean of *ŷ*_*v*_ *− y*_*v*_ and the 95% limits of agreement are the bias *±*1.96 standard deviations of the differences.

Clinical metrics treat reduced EF as the positive class. For each threshold *c ∈ {*40, 50*}*, a video is test-positive when *ŷ*_*v*_ *< c* and condition-positive when *y*_*v*_ *< c*; sensitivity and specificity follow from the resulting confusion matrix, and the area under the receiver operating characteristic curve (AUC) is computed by ranking videos by predicted EF; AUC at both thresholds, with the underlying confusion counts, is listed for every configuration in S1 Table. MAE is also reported within four EF bands (*<*30, 30–40, 40–55, *≥*55; *n* = 83, 77, 241, and 876 test videos respectively). Cost metrics are those of the cost protocol above. No configuration is reported without all three metric families. All per-configuration results underlying the figures and tables are provided in S1 Dataset.

Uncertainty is quantified by the nonparametric bootstrap [34]: 10,000 resamples of the test set drawn with replacement at the video level, which is patient-level resampling because EchoNet-Dynamic contains one video per patient. All intervals are 95% percentile confidence intervals (CIs). The resample indices are generated once from a fixed seed and shared across every configuration, so a difference in any metric between two configurations can be evaluated on identical resamples: for each resample we compute the difference in MAE between the two configurations on the same set of videos, and the two are called distinguishable only when the 95% interval of this paired difference excludes zero. For the deployment question we pre-specified a non-inferiority test [35] against the reference configuration with a margin of 0.5 EF points, roughly one eighth of the reference model’s own error and well below the resolution at which EF is used clinically: a configuration is non-inferior when the upper bound of the paired-difference interval lies below the margin.

Because differences of interest are fractions of a point, the two headline comparisons were additionally replicated under two extra training seeds (20260815 and 20260816), chosen by a fixed calendar rule and pre-registered in the repository before any replicate run started. Four configurations were replicated: the reference, the cheap 16 *×* 4 setting, and the matched pair 8 *×* 1 versus 8 *×* 4 that isolates the sampling period. With three seeds per configuration, each comparison is evaluated across all nine cross-seed pairings, a design that exposes run-to-run training variance that no single-seed comparison can.

The full analysis code, training configurations, per-video predictions, metrics, and cost measurements are available at https://github.com/Kushaan-SSSK/ejection-fraction-on-a-budget.

## Results

Table 1 reports the full cost profile of all 22 configurations, sorted by CPU latency per video, and Table 2 their accuracy and clinical metrics in the same order. The subsections below walk through the sweep axis by axis.

**Table 2.** Accuracy and clinical metrics for every configuration, ordered as in Table 1. MAE with 95% patient-level bootstrap CI; sensitivity and specificity from thresholding the regression output at EF 40% and 50%. ^*†*^X3D caveat as in Table 1.

| Configuration | MAE (95% CI) | RMSE | $R^2$ | Sens<sub>40</sub> | Spec<sub>40</sub> | Sens<sub>50</sub> | Spec<sub>50</sub> |
| --- | --- | --- | --- | --- | --- | --- | --- |
| 2D ResNet-18 + pool 16f×p4 | 5.65 (5.36–5.94) | 7.69 | 0.605 | 0.519 | 0.980 | 0.649 | 0.948 |
| X3D-S 16f×p4<sup>†</sup> | 4.22 (4.01–4.43) | 5.65 | 0.786 | 0.669 | 0.984 | 0.754 | 0.956 |
| 2D ResNet-18 + pool 32f×p2 | 5.64 (5.35–5.95) | 7.85 | 0.588 | 0.456 | 0.992 | 0.537 | 0.972 |
| X3D-M 16f×p4<sup>†</sup> | 4.14 (3.95–4.34) | 5.46 | 0.801 | 0.700 | 0.986 | 0.744 | 0.951 |
| X3D-S 32f×p2<sup>†</sup> | 4.25 (4.04–4.46) | 5.71 | 0.782 | 0.700 | 0.981 | 0.740 | 0.960 |
| R3D-18 16f×p4 | 4.15 (3.94–4.36) | 5.59 | 0.791 | 0.725 | 0.977 | 0.782 | 0.950 |
| R(2+1)D-18 16f×p4 | 4.04 (3.84–4.24) | 5.43 | 0.803 | 0.719 | 0.981 | 0.786 | 0.959 |
| MC3-18 16f×p4 | 4.29 (4.09–4.50) | 5.69 | 0.784 | 0.694 | 0.974 | 0.779 | 0.947 |
| R(2+1)D-18 32f×p4 | 4.07 (3.88–4.28) | 5.48 | 0.799 | 0.700 | 0.980 | 0.733 | 0.953 |
| R(2+1)D-18 8f×p4 | 4.36 (4.14–4.58) | 5.93 | 0.765 | 0.619 | 0.987 | 0.698 | 0.967 |
| X3D-M 32f×p2<sup>†</sup> | 4.23 (4.03–4.44) | 5.62 | 0.789 | 0.650 | 0.983 | 0.740 | 0.950 |
| R3D-18 32f×p2 | 3.99 (3.80–4.19) | 5.37 | 0.807 | 0.713 | 0.987 | 0.754 | 0.966 |
| MC3-18 32f×p2 | 4.28 (4.08–4.49) | 5.67 | 0.785 | 0.694 | 0.986 | 0.761 | 0.958 |
| R(2+1)D-18 32f×p2 (ref.) | 4.04 (3.84–4.24) | 5.45 | 0.801 | 0.725 | 0.979 | 0.775 | 0.947 |
| R(2+1)D-18 64f×p4 | 4.03 (3.84–4.24) | 5.40 | 0.805 | 0.706 | 0.980 | 0.786 | 0.956 |
| R(2+1)D-18 64f×p2 | 4.10 (3.91–4.31) | 5.48 | 0.799 | 0.694 | 0.983 | 0.758 | 0.960 |
| R(2+1)D-18 16f×p2 | 4.20 (4.00–4.41) | 5.62 | 0.788 | 0.681 | 0.987 | 0.737 | 0.961 |
| R(2+1)D-18 8f×p2 | 4.85 (4.61–5.11) | 6.61 | 0.707 | 0.600 | 0.983 | 0.670 | 0.974 |
| R(2+1)D-18 64f×p1 | 4.12 (3.93–4.32) | 5.48 | 0.799 | 0.713 | 0.982 | 0.754 | 0.953 |
| R(2+1)D-18 32f×p1 | 4.16 (3.95–4.37) | 5.64 | 0.787 | 0.613 | 0.988 | 0.723 | 0.973 |
| R(2+1)D-18 16f×p1 | 4.79 (4.54–5.05) | 6.61 | 0.708 | 0.600 | 0.987 | 0.653 | 0.973 |
| R(2+1)D-18 8f×p1 | 5.34 (5.07–5.63) | 7.35 | 0.638 | 0.500 | 0.987 | 0.607 | 0.964 |

### Baseline reproduction

Our reference configuration (R(2+1)D-18, 32 frames, period 2) reached a test MAE of 4.04 (95% CI 3.84–4.24), RMSE of 5.45 (5.14–5.78), *R*^2^ of 0.80 (0.77–0.83), and an AUC of 0.976 (0.966–0.985) for detecting EF below 40% and 0.962 below 50%, against the published 4.1 MAE, 5.3 RMSE, 0.81 *R*^2^, and 0.97 AUC [5], despite the schedule shortened from 45 to 30 epochs. Bland-Altman bias was +0.03 EF points with limits of agreement *−*10.7 to +10.7. Every comparison below is anchored to this run.

### Sparse sampling beats dense sampling

At every fixed frame count, sampling frames further apart improved accuracy (Table 2). At 8 frames, MAE improved from 5.34 (period 1) to 4.36 (period 4); at 16 frames, from 4.79 to 4.04; at 32 frames, from 4.16 to 4.07; and at 64 frames, from 4.12 to 4.03. The mechanism is plain: a period-1 clip of 8 or 16 frames spans a fraction of a cardiac cycle, so the network never sees a full systole-diastole excursion in one forward pass, while period 4 stretches the same frame budget across enough time to cover it. The effect survived seed replication without qualification: across all nine cross-seed pairings of the 8 *×* 1 and 8 *×* 4 configurations, period 4 won every time, by 1.04 MAE points on average (minimum 0.98).

Dense sampling is also the more expensive choice per video, because a short-span clip tiles a video into more clips. The period-1 rows of Table 1 are therefore dominated twice over: worse accuracy at two to nearly five times the per-video cost. Any deployment sampling densely at a small frame count is paying more to do worse.

### Backbones: plain 3D convolutions win

The best accuracy in the entire study came not from the reference R(2+1)D-18 but from plain R3D-18 at the reference clip geometry: MAE 3.99 (95% CI 3.80–4.19), RMSE 5.37, *R*^2^ 0.81, AUC 0.978 at the 40% threshold, at 1122 ms CPU latency per video against the reference’s 1381 ms, a 19% saving (Tables 1 and 2; S1 Table). The paired difference against the reference is *−* 0.045 (95% CI *−* 0.178 to +0.086), statistically indistinguishable. The two backbones are nearly matched in parameters (33.2 versus 31.3 million) and per-clip compute (163.0 versus 162.6 GFLOPs at 32 *×* 2), so the latency gap comes from the factorization itself: splitting each 3D convolution into a spatial and a temporal stage doubles the number of convolution launches and intermediate activations per block, overhead that Table 1 shows never pays for itself in accuracy on this task. This is the opposite of the pattern on large-scale action recognition, where the factorized design was introduced precisely because it outperformed plain 3D convolution at matched capacity [24]. MC3-18, the mixed-convolution design, was slightly but distinguishably worse than the reference at both clip settings (+0.24 to +0.25 MAE) despite a comparable FLOP count, so its 65% parameter saving buys no deployment advantage here either.

The 2D floor collapsed, and the collapse is informative. ResNet-18 with temporal mean pooling, which sees the same frames but cannot model motion, reached only 5.65 MAE at 16 *×* 4 and 5.64 at 32 *×* 2, more than 1.6 points behind the reference, with *R*^2^ falling from 0.80 to 0.59–0.61, AUC at the 40% threshold from 0.976 to 0.938, and sensitivity at that threshold to 0.52 and 0.46 at the two clip settings. EF is a motion quantity, and no amount of per-frame appearance modeling substitutes for temporal structure. This places the compute floor for this task: below roughly 200 ms of CPU time per video, in this study, the only configurations available are ones that have given up temporal modeling, and they are not clinically usable.

X3D landed between those poles, with the caveat described in the Materials and methods. X3D-M at 16 *×* 4 reached 4.14 MAE (indistinguishable from the reference, paired CI *−*0.046 to +0.247) at 490 ms per video, roughly a third of the reference cost, with a 12 MB checkpoint against the reference’s 125 MB. X3D-S at the same geometry reached 4.22 (slightly but distinguishably worse, +0.18) at 223 ms, a sixth of the reference cost. Whether native-resolution decoding would close X3D’s remaining accuracy gap cannot be answered from these data.

### The frontier

Fig 1 shows the full trade space with cost on a log axis, and Fig 2 the same axis against sensitivity at the 40% threshold. Five configurations trace the accuracy frontier: the 2D floor (163 ms, MAE 5.65), X3D-S 16 *×* 4 (223 ms, 4.22), X3D-M 16 *×* 4 (490 ms, 4.14), R(2+1)D-18 16 *×* 4 (688 ms, 4.04), and R3D-18 32 *×* 2 (1122 ms, 3.99). The frontier is steep below half a second and nearly flat above it: the frontier configurations between 688 ms and 1122 ms are statistically indistinguishable from the reference and, for the pair tested, from each other (R(2+1)D-18 16 *×* 4 against R3D-18 32 *×* 2: paired difference +0.042, 95% CI *−*0.090 to +0.172; S1 Dataset), and past 1122 ms additional compute buys nothing at all. The most expensive configuration in the study, at 3267 ms, is less accurate than every other configuration that retains temporal modeling.

**Fig 1.**
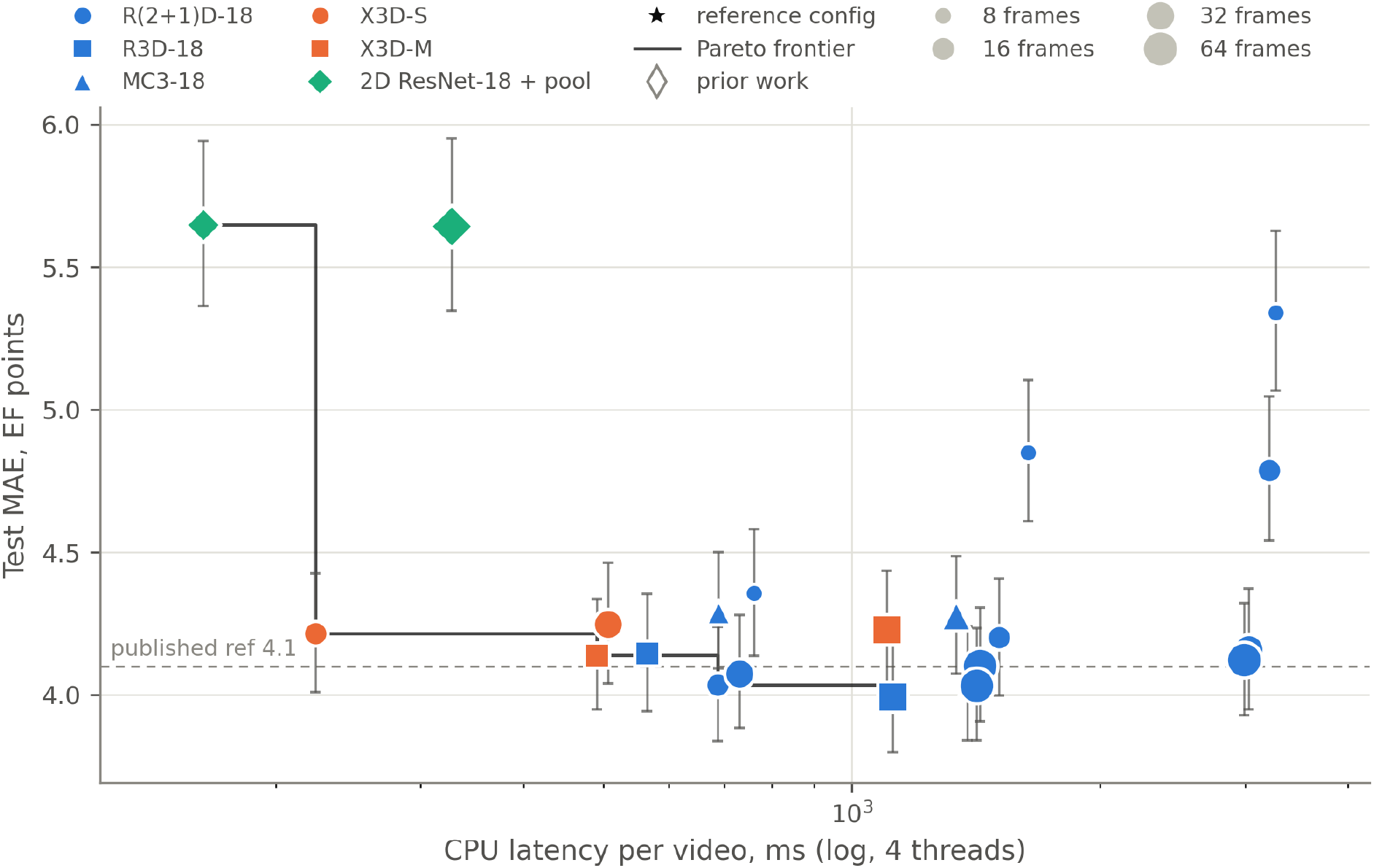
The accuracy-cost trade space. Test MAE (95% bootstrap CI) against CPU latency per video (log axis, 4 threads, median of 100 runs) for all 22 configurations. Color encodes backbone family, marker size clip length, the star the reference configuration, and the stepped line the Pareto frontier. The dashed line marks the published reference MAE of 4.1. X3D points carry the input-resolution caveat described in the Materials and methods.

**Fig 2.**
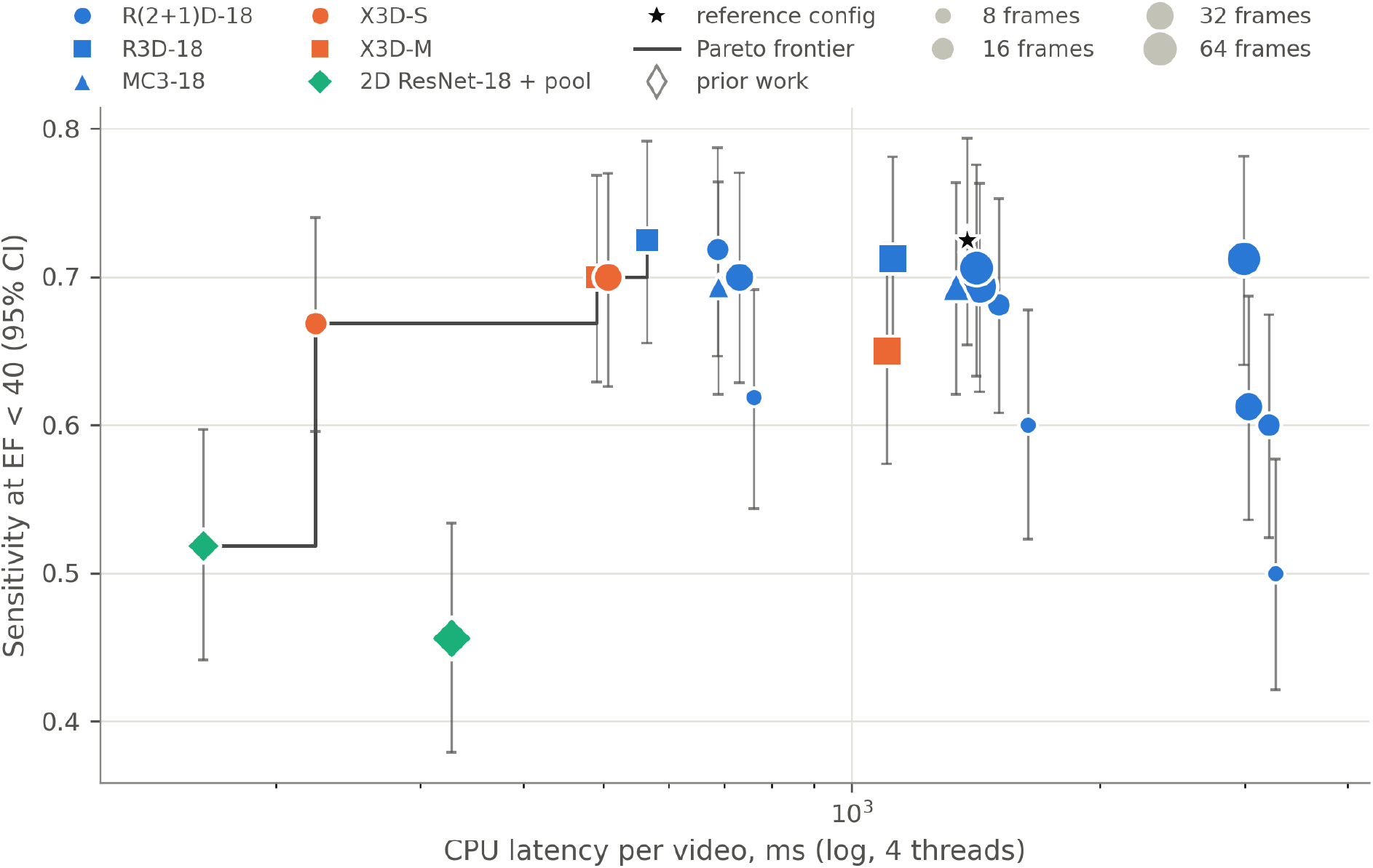
Sensitivity for detecting EF below 40% against cost. Sensitivity (95% bootstrap CI) against the same cost axis as Fig 1. The clinically usable region is confined to configurations that retain temporal modeling.

The frontier’s shape also depends on which cost is counted. Fig 3 redraws the accuracy frontier against FLOPs per video, a hardware-independent measure, and S1 Fig redraws it against GPU latency. The X3D configurations, mid-frontier on latency, dominate the low-cost end on FLOPs: X3D-M at 16 *×* 4 needs 22 GFLOPs per video against the reference’s 371, a 94% reduction, yet delivers only a 65% reduction in measured latency. The discrepancy is the expected behavior of channelwise-separable convolutions, whose low arithmetic intensity (few operations per byte of activation moved) leaves them bound by memory bandwidth rather than by arithmetic throughput [22, 36]; the same effect is well documented for their 2D counterparts [37]. FLOP counts therefore overstate the practical savings of exactly the architecture family they most flatter, which is why measured latency is this study’s primary cost axis.

**Fig 3.**
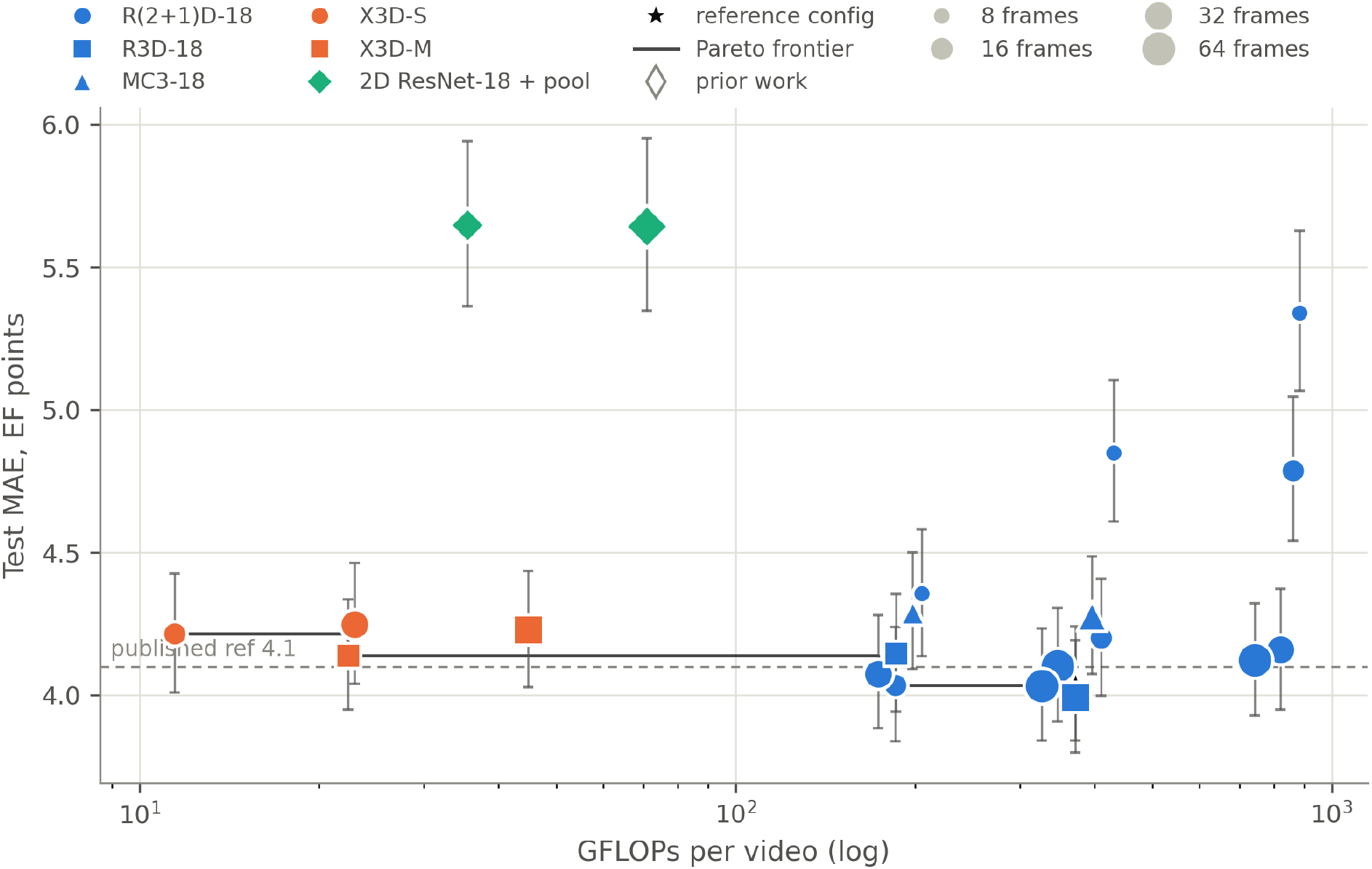
The frontier under a hardware-independent cost. Test MAE (95% bootstrap CI) against FLOPs per video (log axis) for all 22 configurations, with the same encoding as Fig 1. The X3D configurations dominate the low-FLOP region but not the low-latency region.

Sixteen of the 21 non-reference configurations were non-inferior to the reference at the pre-specified 0.5-point margin (Table 3). The cheapest of these with no caveat attached is R3D-18 at 16 *×* 4 (564 ms, 41% of the reference cost); the cheapest overall is X3D-S at 16 *×* 4 (223 ms, 16% of the reference cost), subject to the upsampling confound.

**Table 3.**
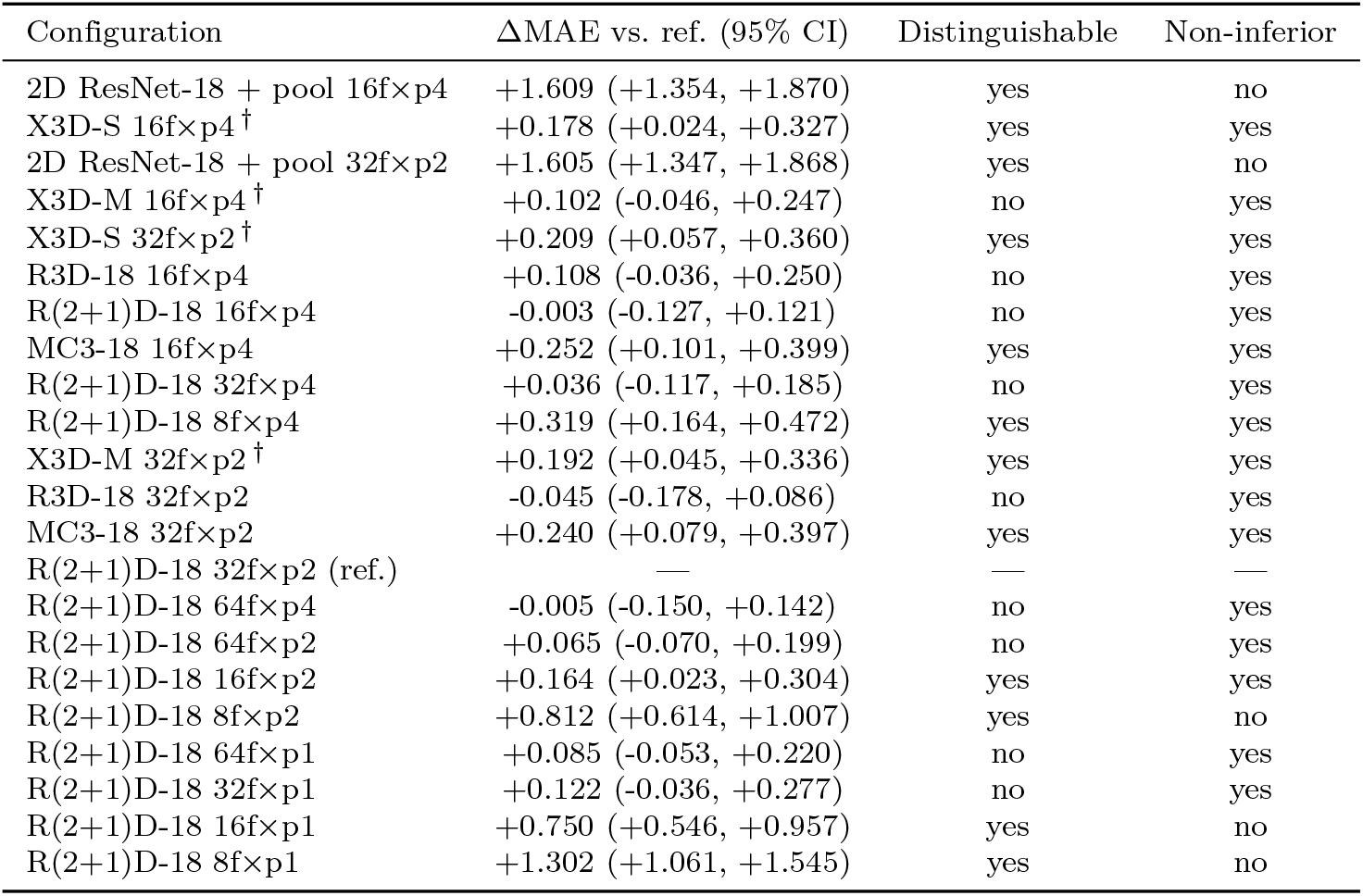
Paired bootstrap comparison of every configuration against the reference. Identical resamples, 10,000 draws. A configuration is distinguishable when the 95% CI of the MAE difference excludes zero, and non-inferior when the upper CI bound is below the pre-specified 0.5-point margin. ^*†*^X3D caveat as in Table 1.

### Clinical error structure

Averages understate what matters clinically, so Fig 4 stratifies error by EF band. Two patterns hold across every configuration. First, error is largest exactly where decisions are made: in the reference configuration, MAE in the 30–40 band (6.7 points) is roughly double that in the *≥*55 band (3.4), and the same two-to-one ratio holds for every 3D backbone. Second, cheap configurations do not degrade uniformly: the 2D floor loses about one point of MAE in the normal band but more than five points in the EF *<*30 band (11.2 versus 5.7 for the reference), so the patients most in need of detection are the ones a too-cheap model misses most; per-band MAE for every configuration is tabulated in S1 Dataset (summary all configs.csv). Sensitivity at the 40% threshold tells the same story more bluntly: 0.71 to 0.73 for the reference and R3D-18, 0.72 for the cheap 16 *×* 4 R(2+1)D, 0.67 for X3D-S, and 0.46 to 0.52 for the 2D floor, at specificities above 0.97 throughout (Table 2). Bland-Altman agreement for the recommended R3D-18 configuration (bias +0.41, limits of agreement *−*10.1 to +10.9) is essentially the same as the reference’s (+0.03, *−*10.7 to +10.7). Fig 5 shows predicted against reference EF for both configurations; agreement is tight in the normal range and spreads below the 40% threshold, consistent with the band-stratified error.

**Fig 4.**
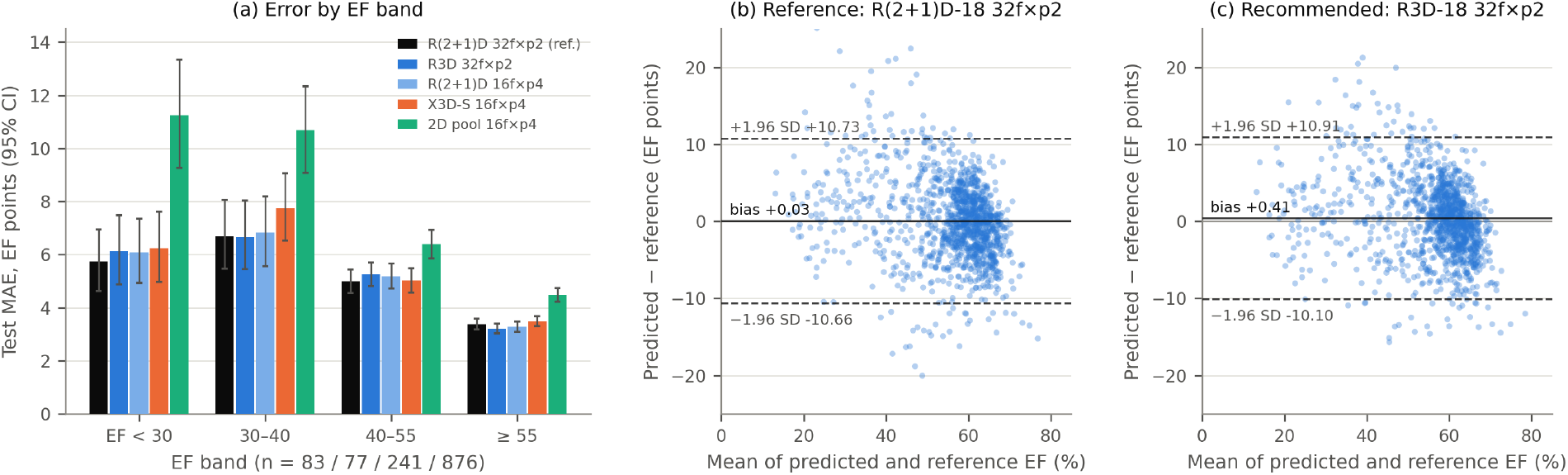
Clinical error structure. A: Test MAE by EF band (95% bootstrap CI) for the reference, the two recommended 3D-CNN configurations (R3D-18 32 *×* 2 and R(2+1)D-18 16 *×* 4), the cheapest non-inferior X3D configuration (X3D-S 16 *×* 4; the recommended X3D-M 16 *×* 4 is not shown), and the 2D floor. Error concentrates in the low-EF bands, and cheap models degrade there first. B and C: Bland-Altman agreement for the reference and the recommended R3D-18 configuration; solid line bias, dashed lines 95% limits of agreement.

**Fig 5.**
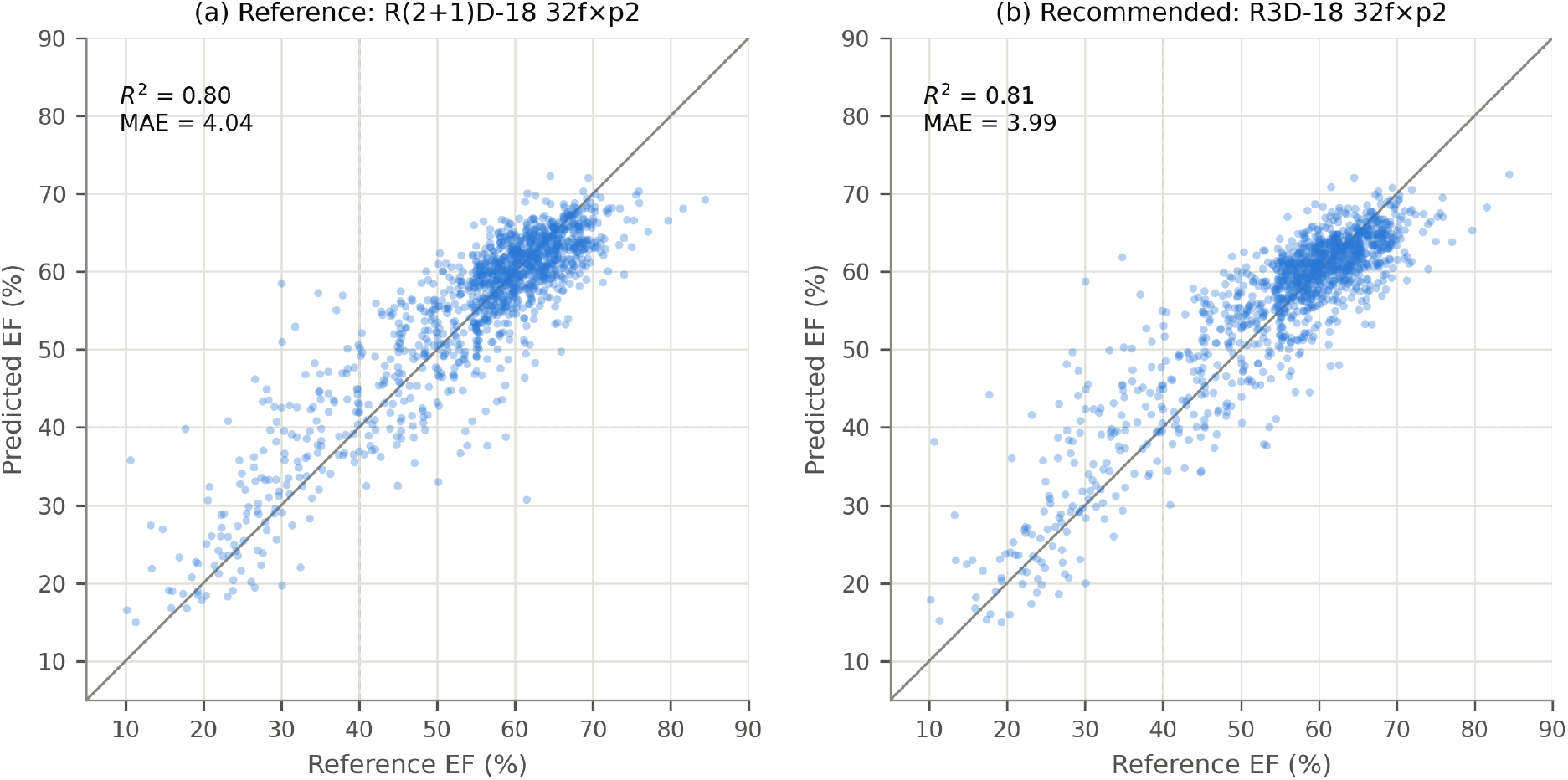
Predicted versus reference EF. A: the reference R(2+1)D-18 32 *×* 2. B: the recommended R3D-18 32 *×* 2. Identity line and the 40% threshold shown; n = 1,277 test videos.

### Seed replication

Table 4 reports the replicate runs. The reference configuration was very stable across seeds (MAE range 0.035), as was 8 *×* 4 (0.026); the cheap 16 *×* 4 setting varied three times as much (0.095), and 8 *×* 1 most of all (0.122).

**Table 4.** Seed replication of the four pre-registered configurations. Seeds 20260814 / 20260815 / 20260816. MAE is shown to three decimals; the ranges, and the cross-seed differences quoted in the text, are computed from the unrounded per-seed values in S1 Dataset (seed variance.csv) and can differ in the last digit from differences of the rounded entries.

| Configuration | MAE by seed | MAE range | Sens<sub>40</sub> by seed |
| --- | --- | --- | --- |
| R(2+1)D-18 32f×p2 (ref.) | 4.038 / 4.030 / 4.003 | 0.035 | 0.725 / 0.675 / 0.688 |
| R(2+1)D-18 16f×p4 | 4.036 / 4.130 / 4.090 | 0.095 | 0.719 / 0.700 / 0.706 |
| R(2+1)D-18 8f×p1 | 5.341 / 5.462 / 5.353 | 0.122 | 0.500 / 0.450 / 0.500 |
| R(2+1)D-18 8f×p4 | 4.357 / 4.331 / 4.337 | 0.026 | 0.619 / 0.631 / 0.644 |

The replication changed one claim and confirmed another, and we report both as measured. The single-seed comparison had found 16 *×* 4 statistically indistinguishable from the reference with a paired difference of *−*0.003. Across the nine cross-seed pairings, however, 16 *×* 4 trailed the reference in eight, by +0.061 MAE on average, about twenty times the single-seed gap. The honest summary is that 16 *×* 4 is approximately tied with the reference, with weak evidence of a small consistent disadvantage that a single seed could not reveal. The deployment conclusion survives (half the cost, no confirmed loss, worst observed cross-seed gap +0.13), but we do not report it as a clean tie. Sensitivity at the 40% threshold showed no such pattern; its cross-seed differences were small and directionless (mean +0.013, spanning *−*0.025 to +0.044). The sparse-versus-dense finding, by contrast, was fully robust, as reported above.

## Discussion

### What to deploy

The study supports three concrete recommendations at three budgets. The budgets are stated as CPU latency at four threads, a deliberate proxy for the application-processor class of hardware found in portable and handheld ultrasound systems [15], though the relative orderings are the more portable quantity. Where the budget accommodates roughly a second of CPU time per video, R3D-18 at 32 *×* 2 is the best choice measured here: the study’s best point accuracy, statistically tied with the reference, at 19% less latency, and a simpler architecture than the reference’s factorized design. Where cost matters more, R(2+1)D-18 at 16 *×* 4 halves the reference’s compute with no statistically confirmed accuracy loss and near-identical sensitivity at the treatment threshold, with the seed-study caveat reported above; R3D-18 at 16 *×* 4 offers a similar deal at 41% of the reference cost. At aggressive budgets, X3D-M at 16 *×* 4 is indistinguishable from the reference in these measurements at a third of the cost, and its 12 MB footprint suits memory-constrained targets, but both X3D results carry the input-resolution confound and should be re-validated with a native-resolution decode before deployment. What no configuration in this study supports is going below the temporal-modeling floor: the 2D-plus-pooling models are 4.2 to 8.5 times cheaper than the reference and clinically unusable, missing roughly half the reduced-EF patients.

The temporal sampling result generalizes beyond any single backbone choice, since it held at every frame count and survived seed replication unqualified: given a fixed frame budget, spread it across at least a full cardiac cycle. Dense short-span sampling is the worst design decision available among configurations that retain temporal modeling: at 8 and 16 frames it degrades accuracy by roughly a full MAE point while multiplying per-video cost, and at 32 and 64 frames the accuracy penalty shrinks to a tenth of a point while the cost multiplier remains.

### Relation to prior work

The original EchoNet-Dynamic paper swept similar axes and observed, as we do, that accuracy saturates with clip length and tolerates coarse sampling [5]; our contribution is attaching measured cost to every such point under one protocol, extending the backbone axis to architectures that postdate 2020, and testing the resulting claims for seed robustness. A recent preprint compares several further video architectures on the same dataset [38], again without a cost dimension. Work proposing individual efficient EF models, such as lightweight segmentation pipelines and compact video transformers [12, 17, 18], is complementary: Echo-E3Net reports 1.55 million parameters and 8.05 GFLOPs [17], and EchoCoTr reports an MAE of 3.95 [12], but each such model is one point in the space this paper maps, and our released protocol allows any of them to be profiled onto the same axes. We did not re-profile prior models here because published implementations target different input pipelines, and cost numbers taken under different conditions are exactly what this study is built to avoid mixing.

The segmentation-based line of work deserves a separate comparison. Pipelines that delineate the ventricle and compute EF geometrically [7–9] offer interpretability that direct regression lacks: a clinician can inspect the contour. Their cost structure is different in kind, since segmentation runs per frame and adds a volume-estimation step, and published segmentation papers on this dataset report accuracy without FLOPs or latency [8, 9], so a fair cost-matched comparison between the two families is an open question our protocol makes answerable. On the deployment side, the edge literature shows that models of the size our frontier’s cheap end occupies do run on single-board hardware in echocardiography [19], and compression techniques such as pruning, quantization, and distillation [20–22] compose with any configuration reported here; our numbers are the uncompressed baselines such techniques would start from. Newer mobile video architectures, MoViNets among them [28], and stronger action-recognition backbones [26, 39] are natural additions to the map as maintained pretrained weights become available.

The R3D-versus-R(2+1)D result also carries a methodological point. On Kinetics-scale action recognition, factorizing spatiotemporal convolutions improves accuracy at matched capacity [24]; on this task the factorized design is more expensive and no more accurate. Echocardiographic EF regression differs from action recognition in ways that plausibly matter: the input is effectively single-channel speckle imagery at 112 *×* 112 rather than natural RGB video, the label is a single continuous quantity driven by one dominant motion pattern rather than one of 400 appearance-rich classes, and the training set is 7,465 videos rather than 240,000 [30]. Whichever of these drives the reversal, the practical lesson is that architecture rankings imported from action recognition do not transfer to this task unexamined, which is an argument for measuring rather than assuming.

### Limitations

The study has clear boundaries. It uses one dataset, one site, one view, and one label convention, at the dataset’s native 112-pixel resolution, so the frontier’s absolute positions should be expected to shift on other distributions even where its shape is likely to transfer. Latency was measured on one GPU model and one CPU configuration; relative orderings are more portable than absolute milliseconds. The cost protocol measures model compute only, excluding video decode and preprocessing, which can be substantial and deployment-specific. The grid shares a single training seed, with seed-variability estimates supplied only for the four seed-replicated configurations; the bootstrap intervals in Tables 2 and 3 capture test-set sampling, not training variance. The seed study itself shows single-seed gaps of a tenth of a point are not trustworthy, which is why no such gap is treated as a ranking in this paper: R3D-18’s *−*0.045 against the reference, in particular, is reported as a tie, not a win. Two cost rows carry the measurement noise described in the Cost protocol. X3D’s numbers carry the upsampling confound throughout. The 0.5-point non-inferiority margin, while pre-specified and small relative to both model error and human interobserver variation [5, 6], is ultimately a judgment; readers who prefer a stricter margin can re-derive the non-inferior set directly from the confidence intervals in Table 3. No quantization, pruning, or distillation was applied [20, 21], and the frontier should be read as a map of architecture and sampling choices, not of every efficiency technique available. The models regress EF directly and were not compared against segmentation-based pipelines under matched cost. Finally, external validation on independent cohorts such as CAMUS [7] or EchoNet-Pediatric [14], and re-measurement of latency on genuine embedded targets in the style of the edge-deployment literature [19], are the natural next steps before any configuration recommended here is built into a device.

## Conclusion

Accuracy for video-based EF estimation on EchoNet-Dynamic is far cheaper than the reference configuration suggests, up to a floor. Half the reference’s compute is available at no statistically confirmed accuracy cost, though with a small seed-consistent disadvantage, a plain R3D-18 matches the reference’s accuracy at 19% less latency, and sparse temporal sampling is strictly better than dense sampling at fixed frame budgets, a result that survived seed replication in all nine pairings. The savings end where temporal modeling ends: models that pool per-frame features miss roughly half the patients below the treatment threshold. The full per-configuration results (S1 Dataset), the cost protocol, and the code are released so that new efficient architectures can be placed on the same map rather than a new one.

## Data Availability

All numerical results underlying the findings (per-video predictions, bootstrapped metrics, cost measurements, and the training-run registry), together with the analysis code and training configurations, are available at https://github.com/Kushaan-SSSK/ejection-fraction-on-a-budget. The EchoNet-Dynamic dataset is available to researchers from Stanford Medicine at https://echonet.github.io/dynamic under a research use agreement that prohibits redistribution; no videos or derived imagery are redistributed by this study.

https://github.com/Kushaan-SSSK/ejection-fraction-on-a-budget

## Supporting information

**S1 Fig. The accuracy frontier against GPU latency**. Test MAE (95% bootstrap CI) against GPU latency per video (log axis, batch size 1, fp32) for all 22 configurations, with the same encoding as Fig 1.

**S1 Table. AUC at the 40% and 50% thresholds for every configuration**. Area under the receiver operating characteristic curve with 95% bootstrap CI, and the true positive, false negative, false positive, and true negative counts at each threshold. (CSV)

**S1 Dataset. Complete numerical results underlying every figure and table**. Per-configuration summary metrics, paired bootstrap comparisons against the reference and best-MAE configurations, the seed replication results, the cost harness output, the training-run registry, per-video predictions, and per-configuration metric files, with a data dictionary. (ZIP)

## Acknowledgments

This research used data provided by the Stanford Center for Artificial Intelligence in Medicine and Imaging (AIMI). AIMI curated a publicly available imaging data repository containing clinical imaging and data from Stanford Health Care, the Stanford Children’s Hospital, the University Healthcare Alliance and Packard Children’s Health Alliance clinics provisioned for research use by the Stanford Medicine Research Data Repository (STARR). We thank the Stanford EchoNet group for making the EchoNet-Dynamic dataset available to the research community.

## References

1. McDonagh TA, Metra M, Adamo M, et al. 2021 ESC Guidelines for the diagnosis and treatment of acute and chronic heart failure. European Heart Journal. 2021;42(36):3599–726. doi:10.1093/eurheartj/ehab368.

2. Heidenreich PA, Bozkurt B, Aguilar D, et al. 2022 AHA/ACC/HFSA Guideline for the Management of Heart Failure. Circulation. 2022;145(18):e895–e1032. doi:10.1161/CIR.0000000000001063.

3. Bozkurt B, Coats AJS, Tsutsui H, et al. Universal Definition and Classification of Heart Failure. Journal of Cardiac Failure. 2021;27(4):387–413. doi:10.1016/j.cardfail.2021.01.022.

4. Lang RM, Badano LP, Mor-Avi V, Afilalo J, Armstrong A, Ernande L, et al. Recommendations for Cardiac Chamber Quantification by Echocardiography in Adults: An Update from the American Society of Echocardiography and the European Association of Cardiovascular Imaging. Journal of the American Society of Echocardiography. 2015;28(1):1–39.e14. doi:10.1016/j.echo.2014.10.003.

5. Ouyang D, He B, Ghorbani A, Yuan N, Ebinger J, Langlotz CP, et al. Video-based AI for beat-to-beat assessment of cardiac function. Nature. 2020;580(7802):252–6. doi:10.1038/s41586-020-2145-8.

6. Pellikka PA, She L, Holly TA, Lin G, Varadarajan P, Pai RG, et al. Variability in Ejection Fraction Measured By Echocardiography, Gated Single-Photon Emission Computed Tomography, and Cardiac Magnetic Resonance in Patients With Coronary Artery Disease and Left Ventricular Dysfunction. JAMA Network Open. 2018;1(4):e181456. doi:10.1001/jamanetworkopen.2018.1456.

7. Leclerc S, Smistad E, Pedrosa J, Ostvik A, Cervenansky F, Espinosa F, et al. Deep Learning for Segmentation Using an Open Large-Scale Dataset in 2D Echocardiography. IEEE Transactions on Medical Imaging. 2019;38(9):2198–210. doi:10.1109/TMI.2019.2900516.

8. Cao K, Zhao M, Geng M, Zheng S, Jung H. Left ventricular segmentation method based on optimized UNet and improved CBAM: ESV and EDV tracking study. PLOS ONE. 2025;20(6):e0325794. doi:10.1371/journal.pone.0325794.

9. Cervantes-Guzmán A, McPherson K, Olveres J, Moreno-García CF, Torres Robles F, Elyan E, et al. Robust cardiac segmentation corrected with heuristics. PLOS ONE. 2023;18(10):e0293560. doi:10.1371/journal.pone.0293560.

10. He B, Kwan AC, Cho JH, Yuan N, Pollick C, Shiota T, et al. Blinded, randomized trial of sonographer versus AI cardiac function assessment. Nature. 2023;616(7957):520–4. doi:10.1038/s41586-023-05947-3.

11. Reynaud H, Vlontzos A, Hou B, Beqiri A, Leeson P, Kainz B. Ultrasound Video Transformers for Cardiac Ejection Fraction Estimation. In: Medical Image Computing and Computer Assisted Intervention – MICCAI 2021. vol. 12906 of Lecture Notes in Computer Science. Springer; 2021. p. 495–505.

12. Muhtaseb R, Yaqub M. EchoCoTr: Estimation of the Left Ventricular Ejection Fraction from Spatiotemporal Echocardiography. In: Medical Image Computing and Computer Assisted Intervention – MICCAI 2022. vol. 13434 of Lecture Notes in Computer Science. Springer; 2022. p. 370–9.

13. Mokhtari M, Tsang T, Abolmaesumi P, Liao R. EchoGNN: Explainable Ejection Fraction Estimation with Graph Neural Networks. In: Medical Image Computing and Computer Assisted Intervention – MICCAI 2022. vol. 13434 of Lecture Notes in Computer Science. Springer; 2022. p. 360–9.

14. Reddy CD, Lopez L, Ouyang D, Zou JY, He B. Video-Based Deep Learning for Automated Assessment of Left Ventricular Ejection Fraction in Pediatric Patients. Journal of the American Society of Echocardiography. 2023;36(5):482–9. doi:10.1016/j.echo.2023.01.015.

15. Chamsi-Pasha MA, Sengupta PP, Zoghbi WA. Handheld Echocardiography: Current State and Future Perspectives. Circulation. 2017;136(22):2178–88. doi:10.1161/CIRCULATIONAHA.117.026622.

16. Narula J, Chandrashekhar Y, Braunwald E. Time to Add a Fifth Pillar to Bedside Physical Examination: Inspection, Palpation, Percussion, Auscultation, and Insonation. JAMA Cardiology. 2018;3(4):346–50. doi:10.1001/jamacardio.2018.0001.

17. Heidari M, Bozorgpour A, Zarif-Fakharnia A, Chen W, Merhof D, Foran DJ, et al. Echo-E3Net: Efficient Endocardial Spatio-Temporal Network for Ejection Fraction Estimation. arXiv preprint arXiv:250317543. 2025. Version 3, cited 2026-08-20; title, author list, and efficiency figures follow this version.

18. Muldoon M, Khan N. Lightweight and Interpretable Left Ventricular Ejection Fraction Estimation Using Mobile U-Net. arXiv preprint arXiv:230407951. 2023.

19. Zhu Y, Gao Y, Wang M, Li M, Wang K. Implementation of resource-efficient fetal echocardiography detection algorithms in edge computing. PLOS ONE. 2024;19(9):e0305250. doi:10.1371/journal.pone.0305250.

20. Han S, Mao H, Dally WJ. Deep Compression: Compressing Deep Neural Networks with Pruning, Trained Quantization and Huffman Coding. In: International Conference on Learning Representations (ICLR); 2016. arXiv:1510.00149.

21. Hinton G, Vinyals O, Dean J. Distilling the Knowledge in a Neural Network. arXiv preprint arXiv:150302531. 2015.

22. Sze V, Chen YH, Yang TJ, Emer JS. Efficient Processing of Deep Neural Networks: A Tutorial and Survey. Proceedings of the IEEE. 2017;105(12):2295–329. doi:10.1109/JPROC.2017.2761740.

23. Feichtenhofer C. X3D: Expanding Architectures for Efficient Video Recognition. In: Proceedings of the IEEE/CVF Conference on Computer Vision and Pattern Recognition (CVPR); 2020. doi:10.1109/CVPR42600.2020.00028.

24. Tran D, Wang H, Torresani L, Ray J, LeCun Y, Paluri M. A Closer Look at Spatiotemporal Convolutions for Action Recognition. In: Proceedings of the IEEE Conference on Computer Vision and Pattern Recognition (CVPR); 2018. p. 6450–9. doi:10.1109/CVPR.2018.00675.

25. Tran D, Bourdev L, Fergus R, Torresani L, Paluri M. Learning Spatiotemporal Features with 3D Convolutional Networks. In: 2015 IEEE International Conference on Computer Vision (ICCV); 2015. doi:10.1109/ICCV.2015.510.

26. Carreira J, Zisserman A. Quo Vadis, Action Recognition? A New Model and the Kinetics Dataset. In: 2017 IEEE Conference on Computer Vision and Pattern Recognition (CVPR); 2017. doi:10.1109/CVPR.2017.502.

27. He K, Zhang X, Ren S, Sun J. Deep Residual Learning for Image Recognition. In: Proceedings of the IEEE Conference on Computer Vision and Pattern Recognition (CVPR); 2016. p. 770–8. doi:10.1109/CVPR.2016.90.

28. Kondratyuk D, Yuan L, Li Y, Zhang L, Tan M, Brown M, et al. MoViNets: Mobile Video Networks for Efficient Video Recognition. In: 2021 IEEE/CVF Conference on Computer Vision and Pattern Recognition (CVPR); 2021. doi:10.1109/CVPR46437.2021.01576.

29. Russakovsky O, Deng J, Su H, Krause J, Satheesh S, Ma S, et al. ImageNet Large Scale Visual Recognition Challenge. International Journal of Computer Vision. 2015;115(3):211–52. doi:10.1007/s11263-015-0816-y.

30. Kay W, Carreira J, Simonyan K, Zhang B, Hillier C, Vijayanarasimhan S, et al. The Kinetics Human Action Video Dataset. arXiv preprint arXiv:170506950. 2017.

31. Paszke A, Gross S, Massa F, Lerer A, Bradbury J, Chanan G, et al. PyTorch: An Imperative Style, High-Performance Deep Learning Library. In: Advances in Neural Information Processing Systems 32 (NeurIPS); 2019. p. 8024–35.

32. FAIR (Meta AI). fvcore: Collection of common code for FAIR computer vision research; 2023. https://github.com/facebookresearch/fvcore.

33. Bland JM, Altman DG. Statistical Methods for Assessing Agreement Between Two Methods of Clinical Measurement. The Lancet. 1986;327(8476):307–10. doi:10.1016/S0140-6736(86)90837-8.

34. Efron B. Bootstrap Methods: Another Look at the Jackknife. The Annals of Statistics. 1979;7(1):1–26. doi:10.1214/aos/1176344552.

35. Walker E, Nowacki AS. Understanding Equivalence and Noninferiority Testing. Journal of General Internal Medicine. 2011;26(2):192–6. doi:10.1007/s11606-010-1513-8.

36. Williams S, Waterman A, Patterson D. Roofline: An Insightful Visual Performance Model for Multicore Architectures. Communications of the ACM. 2009;52(4):65–76. doi:10.1145/1498765.1498785.

37. Howard AG, Zhu M, Chen B, Kalenichenko D, Wang W, Weyand T, et al. MobileNets: Efficient Convolutional Neural Networks for Mobile Vision Applications. arXiv preprint arXiv:170404861. 2017.

38. Saranyan S, Saha P. Investigating Deep Learning Models for Ejection Fraction Estimation from Echocardiography Videos; 2025. ArXiv preprint arXiv:2512.22657.

39. Feichtenhofer C, Fan H, Malik J, He K. SlowFast Networks for Video Recognition. In: 2019 IEEE/CVF International Conference on Computer Vision (ICCV); 2019. doi:10.1109/ICCV.2019.00630.

