## Supplementary material for "Ejection fraction on a budget: mapping the accuracy-compute trade space for video-based ejection fraction estimation": S1 Dataset: README.txt

S1 Dataset: Ejection fraction on a budget
==========================================

Complete numerical results underlying every figure and table in the
manuscript. All EF values are in percent; errors are in EF points;
latencies are in milliseconds; memory and disk sizes in MB.

FILES
-----

summary_all_configs.csv
  One row per primary configuration (22 rows). Accuracy (mae, rmse, r2,
  with 95% bootstrap CI columns *_ci_lo/*_ci_hi), Bland-Altman bias and
  limits of agreement, sensitivity/specificity at the EF 40 and 50 cut
  points, MAE and n per EF band (mae_band_<lo>_<hi>, n_band_<lo>_<hi>;
  bands are lower-bound inclusive), and the frozen-protocol cost columns
  (gflops_per_clip, params_total, disk_mb, gpu/cpu latency per clip and
  per video, gpu_peak_mem_mb). The caveat column flags the X3D
  input-upsampling confound described in the manuscript.

paired_comparisons.csv
  Paired bootstrap MAE differences of every configuration against the
  reference (3a_r2p1d_f32_p2) and against the best-MAE configuration
  (3b_r3d_f32_p2), on shared resample indices; 95% CI bounds and the
  distinguishable flags (CI excludes zero).

seed_variance.csv
  The seed-replication study: test MAE and sensitivity at EF < 40 for
  each of the 4 pre-registered configurations under seeds 20260814,
  20260815, 20260816.

costs.csv
  The cost harness output, one row per configuration: measurement
  protocol tag, input shape, clips per test video, parameters, disk
  size, GFLOPs per clip (2 x multiply-accumulates; unsupported ops
  listed, never dropped), CPU latency (4 threads, median and IQR of 100
  timed runs after 20 warmup runs), GPU latency and peak memory, and the
  derived per-video columns.

registry.csv
  One row per training run (30 runs): timestamp, config, seed, epochs,
  best validation MAE, training hours, device, mixed-precision flag,
  and any per-run deviations (gradient accumulation factor; X3D internal
  upsampling).

predictions/<run>.pred.csv
  Per-video test-set predictions for each of the 30 training runs:
  video file name, reference EF (ef_true), predicted EF (ef_pred), and
  the number of non-overlapping clips averaged (n_clips) under the
  study's tiling protocol.

metrics/<run>.metrics.json
  Full bootstrapped metrics for each run (10,000 patient-level
  resamples): point estimates and 95% percentile CIs for all accuracy
  and clinical metrics reported in the manuscript.

The analysis code that produced these files is available at
https://github.com/Kushaan-SSSK/ejection-fraction-on-a-budget
