## Supplementary figures and images for "Ejection fraction on a budget: mapping the accuracy-compute trade space for video-based ejection fraction estimation"

### S1 Figure

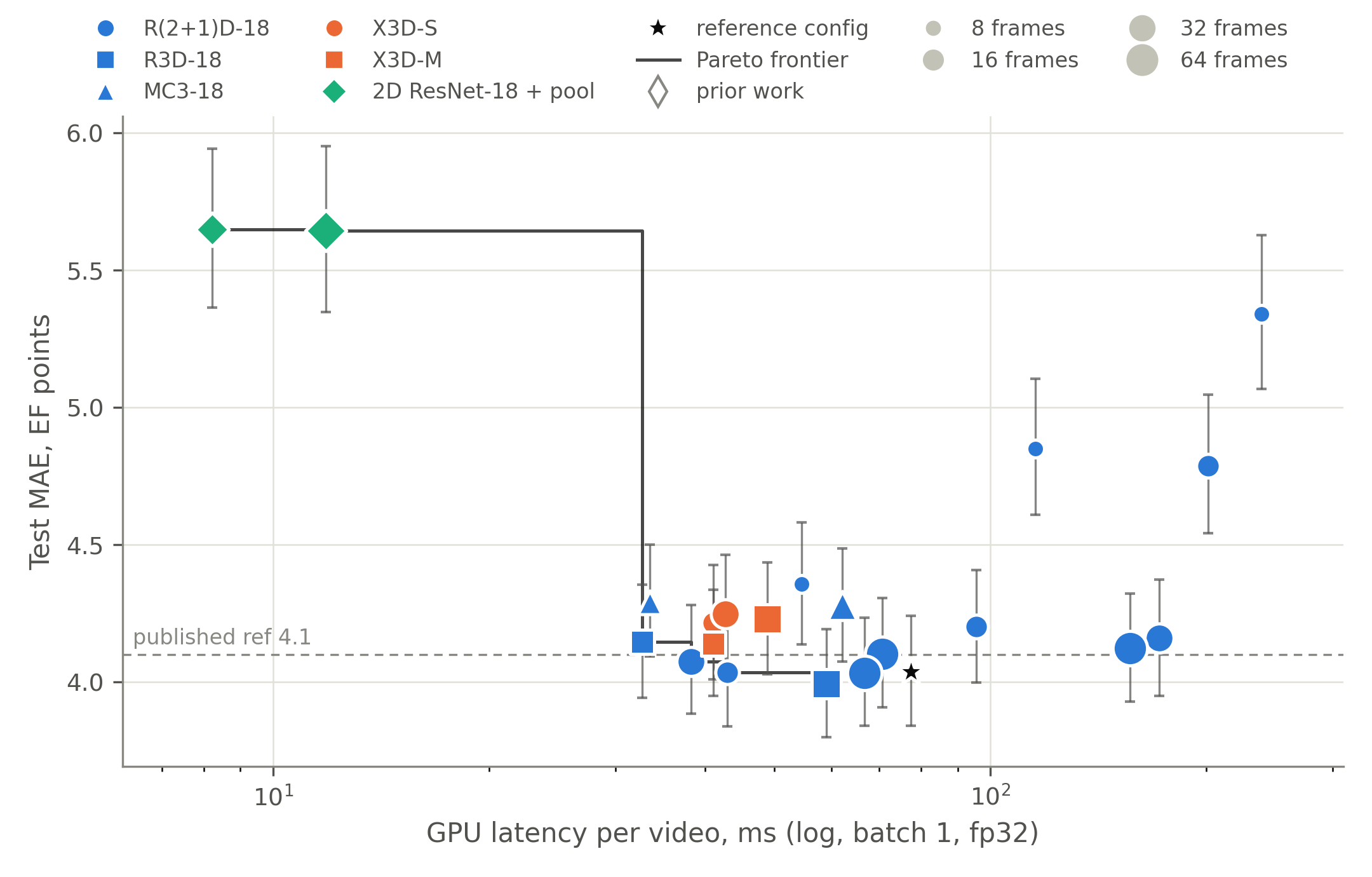
